# Impact of COVID-19 on lifestyle and mental wellbeing in a drought-affected rural Australian population: A mixed method approach

**DOI:** 10.1101/2022.09.23.22280278

**Authors:** Jack Carlson, Kevin Chan, Jonah Gray, Houston Xue, Krista Reed, Jannine K Bailey, Tegan Dutton, Uchechukwu Levi Osuagwu, Robyn Vines

## Abstract

**Background:** The Coronavirus disease 2019 (COVID-19) pandemic has caused unprecedented social and economic disruption, accompanied by the enactment of a multitude of public health measures to restrain disease transmission. These public health and social measures have had a considerable impact on lifestyle and mental wellbeing, which has been well-studied in metropolitan populations, but very little in rural populations. Additionally, the development and use of a standardised scoring system for an overall assessment of patient lifestyle management, and monitoring of changes in these, may be warranted in clinical practice.

**Methods:** The associations between psychological distress and changes in SNAPS health behaviours (smoking, nutrition, alcohol, physical activity, sleep) since the onset of COVID-19 in rural Australia were examined. A cross-sectional anonymous online survey was distributed among adults in the Western New South Wales Primary Health Network in August 2020. The survey included measures of psychological distress, income, disposition, lifestyle factors and behaviours during the pandemic, as well as changes in lifestyle due to COVID-19. A novel Global Lifestyle Score (GLS) was generated as a holistic assessment of lifestyle across multiple domains.

**Results:** The survey was completed by 308 individuals (modal age group: 45-54 years old, 86.4% female). High distress on the K5 scale was present in over one-third of respondents (n=98, 34.3%). Negative change was reported for sleep (24.4%), nutrition (14.3%), alcohol (17.8%), physical exercise (33.8%) and smoking (26.6%) since the onset of the pandemic. Additionally, changes in sleep, nutrition, physical activity and smoking were associated with distress. Respondents with a poor lifestyle (GLS) during the pandemic were significantly more distressed. Perceived COVID-19 impact was associated with high distress, level of drought impact and loss of income.

**Conclusion:** High rates of distress amongst rural Australians during the COVID-19 pandemic was linked, worsening lifestyles as measured by the GLS and loss of income. Lifestyle promotion strategies should be considered by health professionals for the management of crisis-related distress. Further research may explore the impact of COVID-19 on a larger population, including a greater proportion of male respondents, and the impact of modifying lifestyle factors on the reduction of distress in the context of a stressor such as this pandemic.

## Introduction

According to the World Health Organisation (WHO), as of 1 May 2021, the Coronavirus disease 2019 (COVID-19) had infected over 150 million people and been responsible for over 3 million deaths worldwide. Since then, these numbers have more than doubled, reaching 605 million infected people and 6.5 million deaths worldwide(1). In response to the relatively high mortality and morbidity of the pandemic, governments internationally enacted a myriad public health measures, including mandatory social distancing, mask wearing, self-isolation, quarantine and lockdowns(2). At the first peak in April 2020, over half of the global population was under lockdown. Fear, uncertainty, disruption of social interaction and closure of businesses, schools and recreational facilities have had extensive health, economic and social impacts, with the scale of the global economic contraction comparable to the Great Depression of the 1930s(3). The COVID-19 pandemic has been associated with poorer mental health and higher psychological distress in various populations, globally(4, 5).

One area requiring further investigation is the impact this changing physical and social environment has had on lifestyle behaviours - smoking, nutrition, consumption of alcohol, physical activity, and sleep (SNAPS). It is well known that these highly-modifiable lifestyle behaviours are bi-directionally linked to mental health; this likely remains true in the time of COVID-19(6, 7). However, there is a poor understanding of how COVID-19 may have affected these lifestyle behaviours and the relationship between lifestyle and mental wellbeing in this context. An increased understanding of this could provide clinicians and public health organisations with the confidence to target specific behaviours in the prevention and treatment of pandemic-related mental health issues, post COVID-19.

An Australian cross-sectional study of an urban population reported decreased physical activity (48.9% of the sample of 1491 adults) and sleep (40.7%), alongside increased smoking (6.9%) and alcohol consumption (26.6%), during the pandemic(7). These changes were associated with higher depression, anxiety and stress related symptoms, especially negative change in self-reported sleep quality, which had the strongest correlation with depression out of all lifestyle factors examined(8). Similar cross sectional studies from Croatia and the United Kingdom reported strong correlations between poorer sleep, diet and exercise and negative mood, but not with alcohol consumption(9). However, these studies were limited to urban populations and some had incomplete assessment of lifestyle behaviours.

Rural Australians’ experience of COVID-19 is not yet well-represented in the literature. Broadly, the issue of mental illness in rural Australia is exemplified by a high suicide rate, which is up to 40% higher than that of urban areas(10). Rural Australians face a range of general barriers to engagement with mental health services, such as increased physical distance and transport, reduced service availability, a culture of self-reliance and reluctance to discuss mental health issues, in part due to perceived stigma and reduced anonymity(11, 12). In contrast, rural areas benefit from high levels of ‘social capital’, with high rates of community participation and levels of support(13). It remains unknown how living rurally may modify the mental health symptoms in the context of the COVID-19 pandemic. Australian research has demonstrated the development of detrimental health behaviours among rural women during the COVID-19 pandemic, with one study finding that women with children were more likely to report higher high-risk alcohol intake(14).

Economic prosperity has been found to correlate strongly with distress, with loss of income strongly associated with the development of mood and substance use disorders(15). It is suspected that those with a loss of income are the most affected by COVID-19 and have the highest distress. In addition, in recent years, the rural New South Wales population has experienced a drought crisis, with the Millennium Drought lasting between 2001-2009 and the recent drought subsiding only in early 2020 as the COVID-19 pandemic began(16, 17). It is known that drought substantially increases psychological distress and risk of suicide among certain groups, especially males(18). However, it is not known how drought affects lifestyle behaviours. Framing this study in the context of both drought and COVID-19 may improve our understanding of patterns of lifestyle behavioural change in response to these two adversities. This study explored the relationship between lifestyle behaviours and psychological distress in a drought-affected rural Australian region, after the onset of and during the COVID-19 pandemic. There is a paucity of research involving the use of a clinically applicable lifestyle scoring system. Through this study, a ‘Global Lifestyle Score’ (GLS) was created in an attempt to correlate multiple lifestyle factors with the impact of COVID-19 and distress. A holistic and standardised measurement of lifestyle parameters could be applicable in multiple healthcare settings and is in line with current RACGP guidelines requiring comprehensive lifestyle factor screening in patients by their GPs. This study sought to answer the following research questions:

1. What has been the effect of COVID-19 on lifestyle behaviours and income?
2. What is the association between psychological distress - in the context of COVID-19 - and current lifestyle behaviours, lifestyle change, income disruption and disposition?
3. What is the association between respondents reported perceived impact of COVID-19, and current lifestyle behaviours, lifestyle change, income change and disposition?
4. Does a novel composite ‘Global Lifestyle Score’ have utility in predicting known correlates of lifestyle behaviours, including psychological distress?
5. Is there a correlation between respondents’ perception of the impact of the separate crises of COVID-19 and the recent drought?

## Methods

### Study design and setting

Cross-sectional study in the Western New South Wales (NSW) Primary Health Network Region.

### Survey respondents

An anonymous, online survey was conducted in August 2020 and distributed to various rural communities via Facebook and local email mailing lists. Eligible respondents were any adults aged 18 and over currently living in the study region. At the time of distribution, Victoria was in the midst of a stage 4 lockdown with various unlinked “mystery” cases appearing in NSW prompting significant social distancing, partial lockdown and travel restrictions. Social distancing measures included keeping a minimum 1.5 meters between people and strict limitations to public gatherings. Many restaurants, bars and retail stores had strict capacity limits based on their ability to maintain social distancing. Most schools were slowly returning to in-person study after months of online study. University campuses limited or ceased face-to-face teaching and transitioned to online learning, with most clinical placements postponed or cancelled. Most states had closed their borders to NSW and travel to regional areas was not encouraged.

### Survey Tool

The questions for the survey were developed around sleep and the SNAP lifestyle guidelines of the RACGP(18). To assess mental wellbeing, the Kessler-5 (K5) and Adult Dispositional Hope Score (ADHS) were incorporated into this survey. Additional questions around COVID-19 impact, drought impact and demographic information including age, gender, occupation, and postcode were collected. Further detail of the survey structure is shown in the supplementary file (S1 File). Due to COVID-19 and the difficulty in distributing surveys in-person, community engagement was primarily achieved online, by contacting local councils, who distributed a survey link via email and by posting on community Facebook groups for residents of Central West NSW. The survey was accessible on mobile devices and computers.

### Assessing the impact of COVID-19 on the lives of respondents and mental welfare

Details of the response structure are also available in the supplementary file (S1 File). Briefly, the impact of COVID-19 on income was measured through estimate of hours worked per week using a Likert scale. Information on government COVID-19 payments was also obtained. For subjective assessment of the impact of COVID-19, respondents were asked, “Overall, the COVID-19 situation has had a negative impact on my life, my mental health and wellbeing, my financial situation, my work, my ability to provide for myself and my family”. Physical exercise was reported as the number of days per week doing exercise, classified into moderate intensity exercise for greater than 30 minutes, high intensity exercise for greater than 30 minutes and muscle strengthening exercise. These categories conform to the Australian lifestyle recommendations, which are based on the Metabolic Equivalent Task (MET) measurements for physical activities(19). These exercise categories were collated using by calculating and using the MET scores(20).

Sleep was assessed by asking respondents how many hours they slept on average and if the COVID-19 pandemic had changed their sleeping habits. For smoking behaviour, respondents were asked whether they had ever been a regular smoker and/or a current smoker. The change in number of cigarettes smoked daily since the onset of the COVID pandemic was assessed, as was the change in number of stand drinks consumed using the Alcohol Use Disorder Identification Test Consumption (AUDIT-C)(21). The usual number of standard drinks consumed and episodes consuming more than 6 standard drinks was surveyed. Respondents were then asked if the COVID-19 Pandemic had changed their drinking habits

Current nutrition was assessed using a sliding scale for both the number of vegetables and fruits consumed, based on current government recommendations for five vegetable and two fruit servings a day(22).

### Global Lifestyle Score (GLS)

A GLS for each respondent was created by grading reported behaviour for each of the five lifestyle items (smoking, nutrition, alcohol, physical activity, and sleep) against Group consensus guidelines as shown in Table 1. Composite score was calculated and ranged from 5 to 15, with higher scores up to a ceiling of 15 indicating a healthier overall lifestyle profile (Table 1).

**Table 1:**
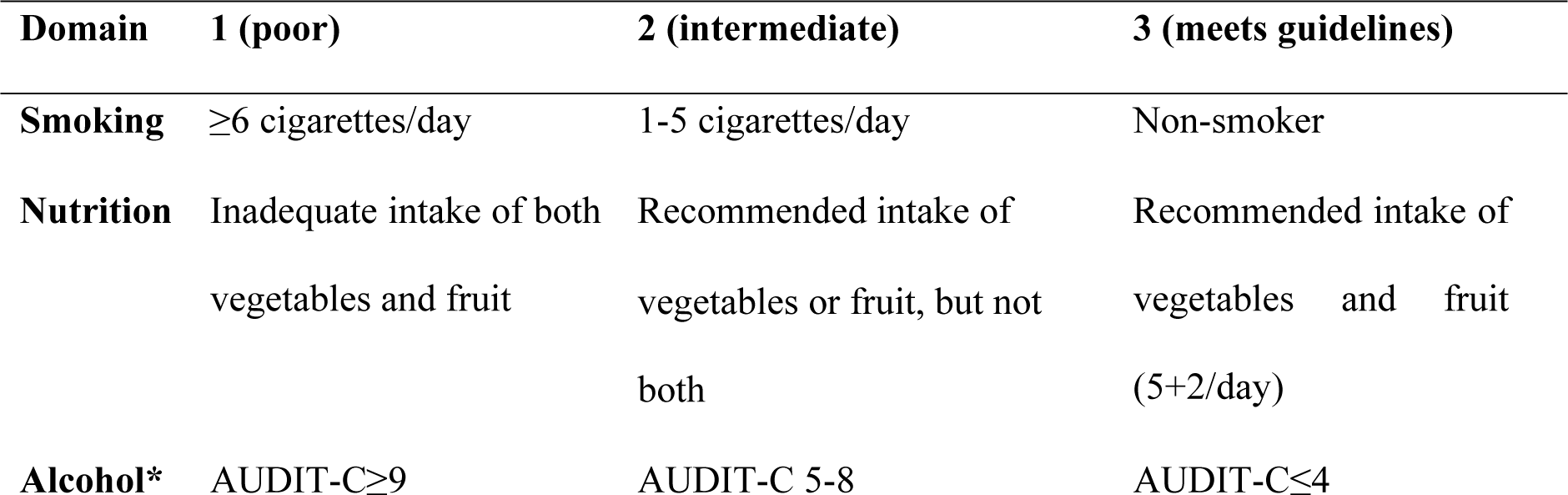
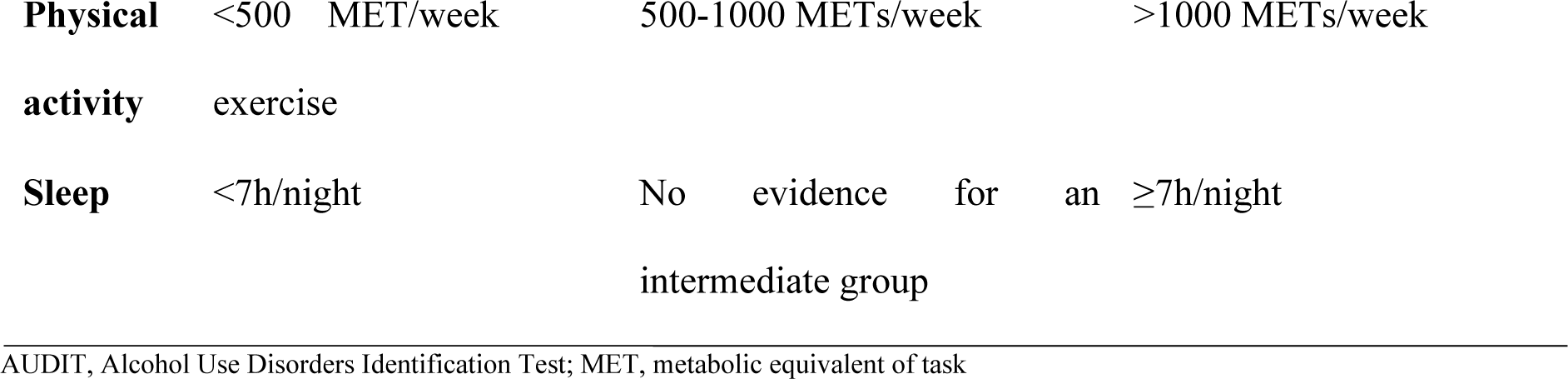
Global Lifestyle Score (GLS) Scoring Rubric.

### Mental health and wellbeing measures

To assess mental wellbeing, the Kessler-5 (K5) for psychological distress and the Adult Hope Scale (AHS: a standardised score calculated from addition of agency and pathway subscales on the Adult Hope Scale) for hopeful dispositional traits were incorporated into this survey. The validated K-5 questionnaire has 5 items which are designed on a Likert scale with the scores ranging from ‘0’ (never) to 5 (all the time)(23). The Adult Hope Scale (AHS)(24), uses 12 statements such as “My past experiences have prepared me well for the future” with responses designed on an 8 point scale. The scores ranged from 1 for “Definitely untrue” to 8 for “Definitely true”. A Total Hope Score (THS) was calculated by adding the Pathways Subscale Score (the sum of items 1, 4, 6 and 8) and Agency Subscale Score (the sum of items 2, 9, 10 and 12), giving a range of scores from 8 to 64, with higher scores representing higher hope levels.

### The impact of drought

The impact of the drought on this rural population was assessed using the following question: “Prior to COVID, how was the drought affecting the following components of your lifestyle?”. A 5-item Likert scale that included impact on sleep, nutrition, alcohol, physical exercise and smoking was used, with scores ranging from 1 for ‘much better’ to 5 for ‘much worse’

### Data Analysis

Statistical analysis was performed on IBM SPSS Version 27 (SPSS Inc., Armonk, NY). For analysis, Likert scale responses for overall COVID-19 impact, drought impact and impact of COVID-19 on income, smoking, nutrition, alcohol, physical activity, and sleep were recoded into subcategories of negative impact (1), neutral (2), or positive impact (3), to maximise power. Results were presented using descriptive statistics including frequencies and percentages for categorical variables (e.g., demographic variables and COVID-19 impact scales), and means ± standard deviations (SD) for continuous variables. Independent sample *t*-test or ANOVA with *post-hoc* Tukey tests where necessary, were used to compare the K-5 and the THS between groups, based on the demographics, overall COVID-19 impact and COVID-19 income impact. Age was recategorized into <35, 35-54 or >54 years, age groups. The Pearson correlation coefficient was used to assess the relationship between K5 scores and the GLS and THS. The associations between overall COVID-19 impact and demographics and reported impact of COVID-19 on individual lifestyle factors were determined using Chi-square tests. Appropriate statistics (Pearson’s r, t, F values, df) were reported for all tests. All tests were two-tailed and *P*<0.05 was considered statistically significant.

Respondents’ qualitative comments were analysed by brief thematic analysis to identify key themes regarding the impact of the COVID-19 pandemic on lifestyle.

## Results

### Characteristics of the respondents

The sociodemographic characteristics of respondents are reported in Table 2 and in Figures 1, 2 and 3. Most respondents were females (86.4%), aged between 45-54 years old (27.6%) and were working in a non-agricultural industry (76.2%) at the time of this study. For the mood and disposition measures, over one-third of respondents (34.3%) reported symptoms of psychological distress and the mean AHS was 44.1 ±10.7.

**Table 2:**
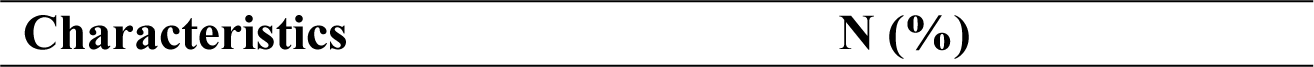
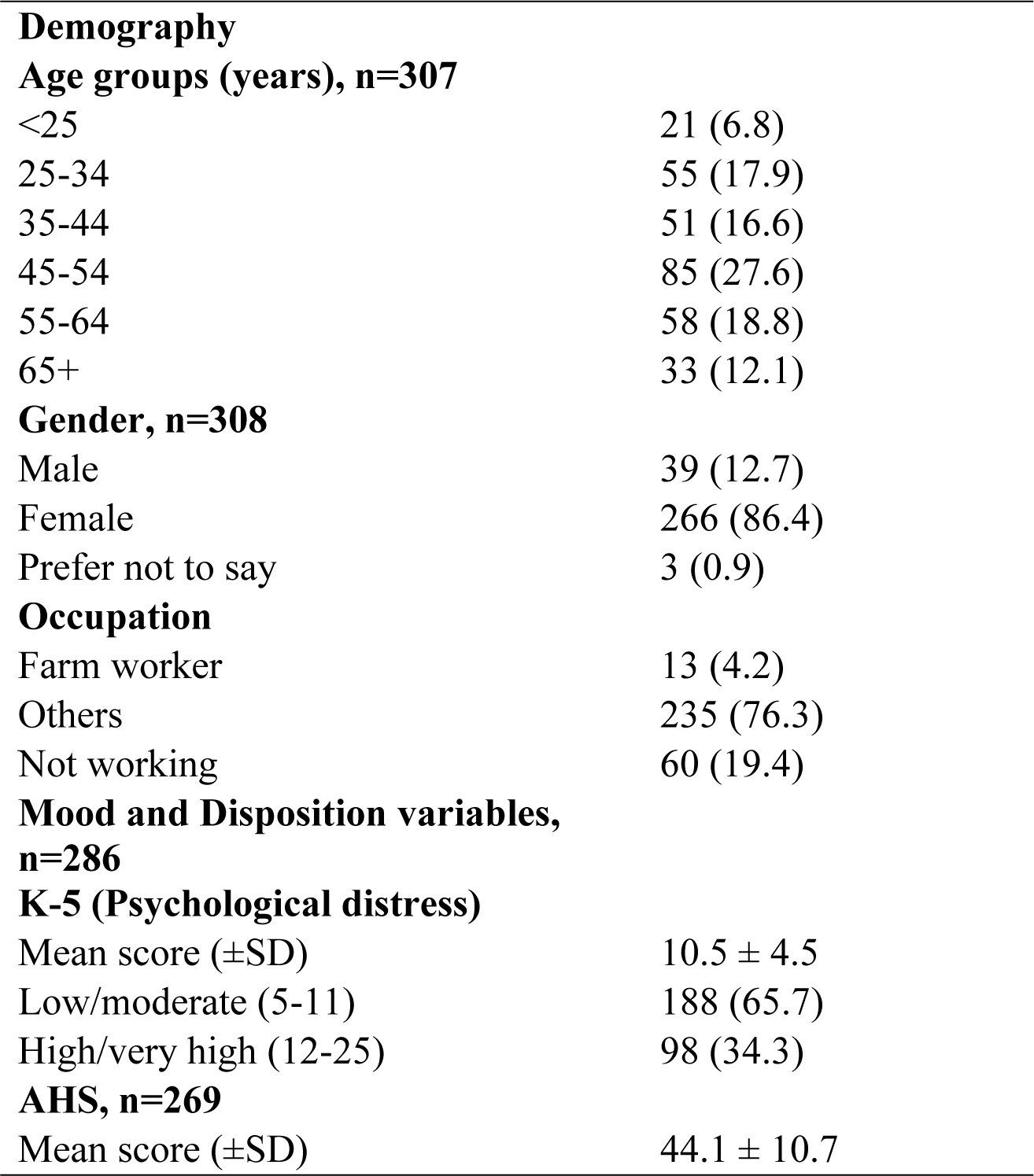
Characteristics of the respondents and the health scores.

| Characteristics | N (%) |
| --- | --- |
| **Demography** |  |
| **Age groups (years), n=307** |  |
| <25 | 21 (6.8) |
| 25-34 | 55 (17.9) |
| 35-44 | 51 (16.6) |
| 45-54 | 85 (27.6) |
| 55-64 | 58 (18.8) |
| 65+ | 33 (12.1) |
| **Gender, n=308** |  |
| Male | 39 (12.7) |
| Female | 266 (86.4) |
| Prefer not to say | 3 (0.9) |
| **Occupation** |  |
| Farm worker | 13 (4.2) |
| Others | 235 (76.3) |
| Not working | 60 (19.4) |
| **Mood and Disposition variables, n=286** |  |
| **K-5 (Psychological distress)** |  |
| Mean score (±SD) | 10.5 ± 4.5 |
| Low/moderate (5-11) | 188 (65.7) |
| High/very high (12-25) | 98 (34.3) |
| **AHS, n=269** |  |
| Mean score (±SD) | 44.1 ± 10.7 |

**Figure 1:**
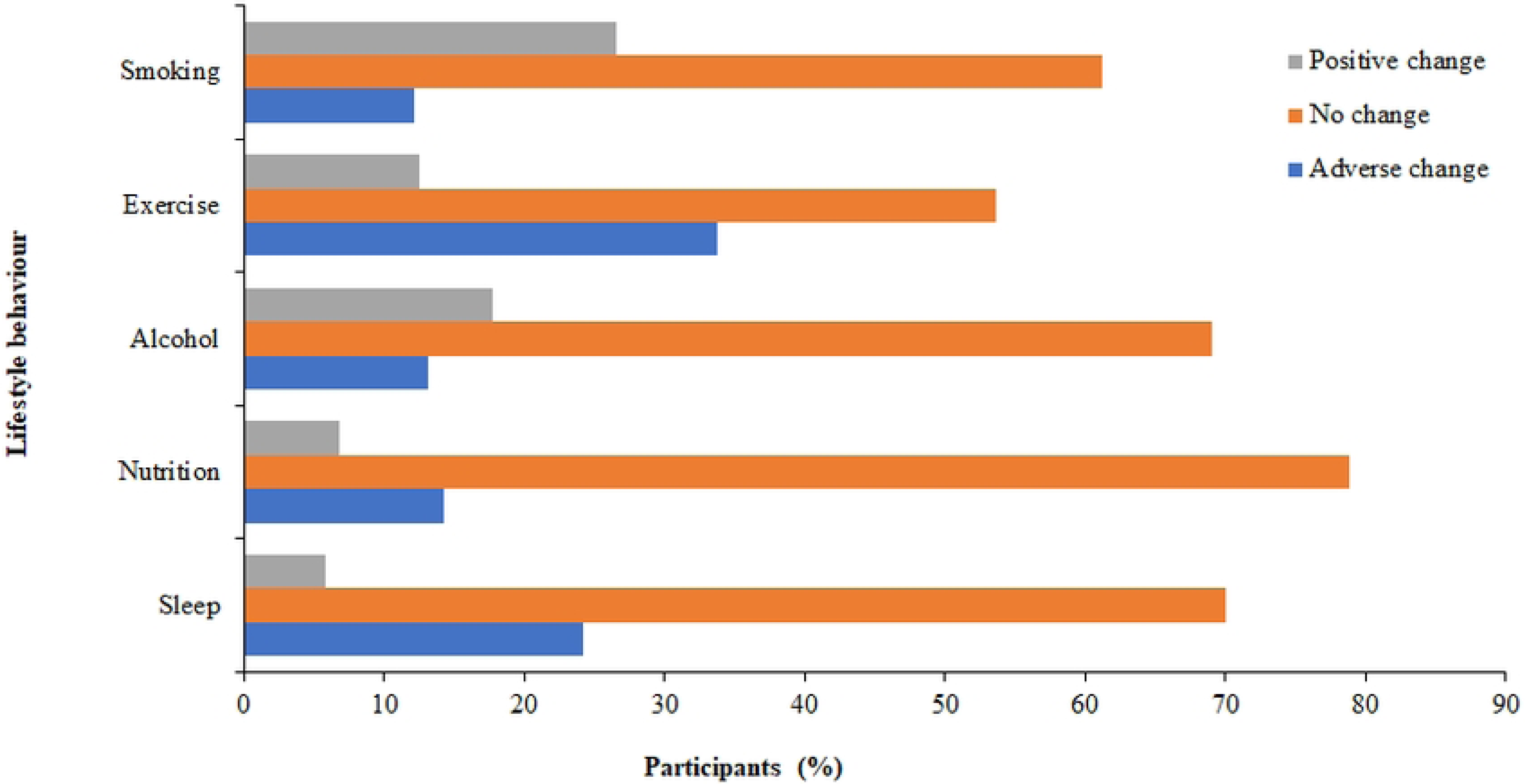
Self-reported Changes in Lifestyle Behaviour due to COVID-19.

**Figure 2:**
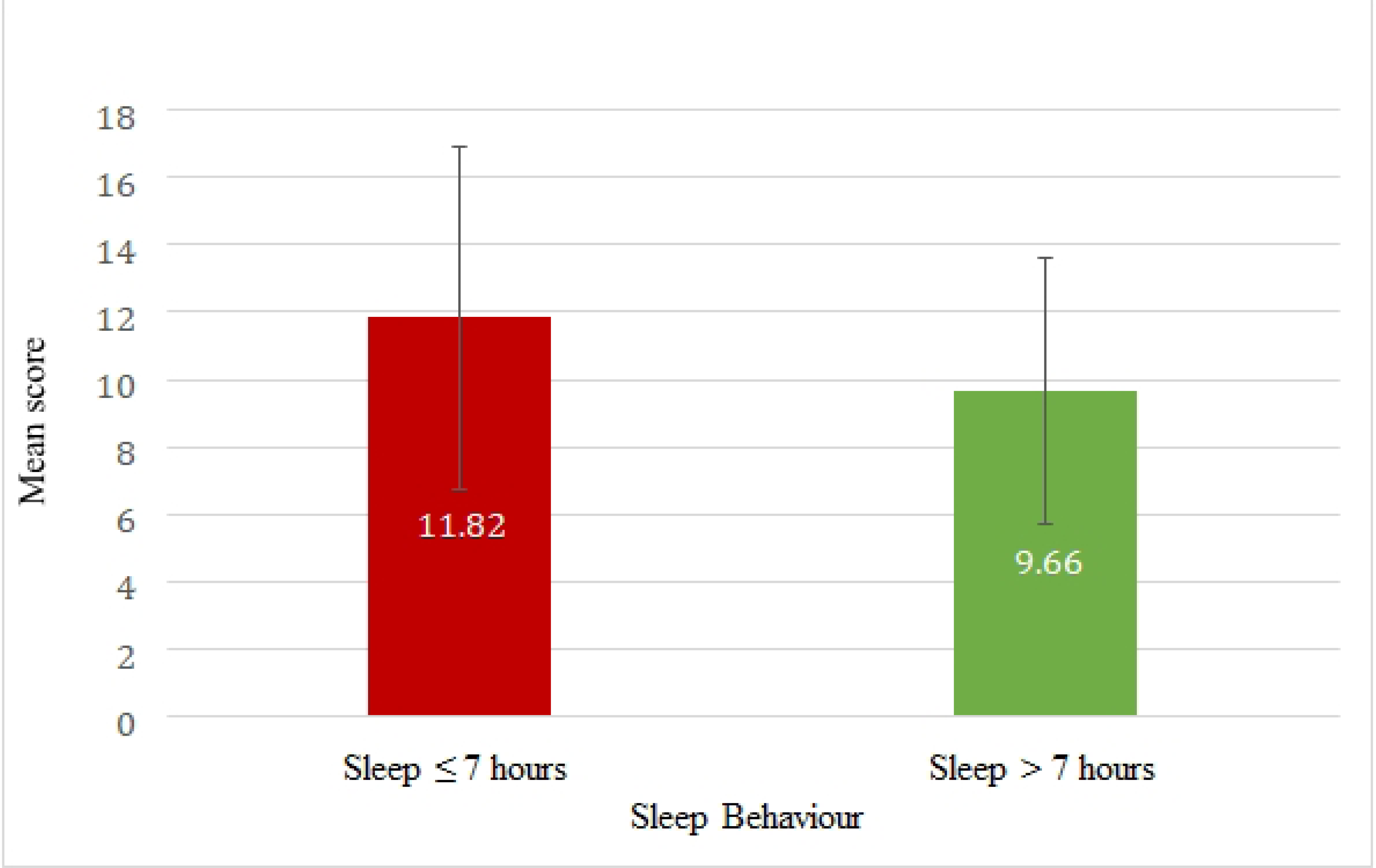
Mean K-5 scores versus the hours of sleep per night due to COVID-19 pandemic. *Error bars are shown*.

**Figure 3:**
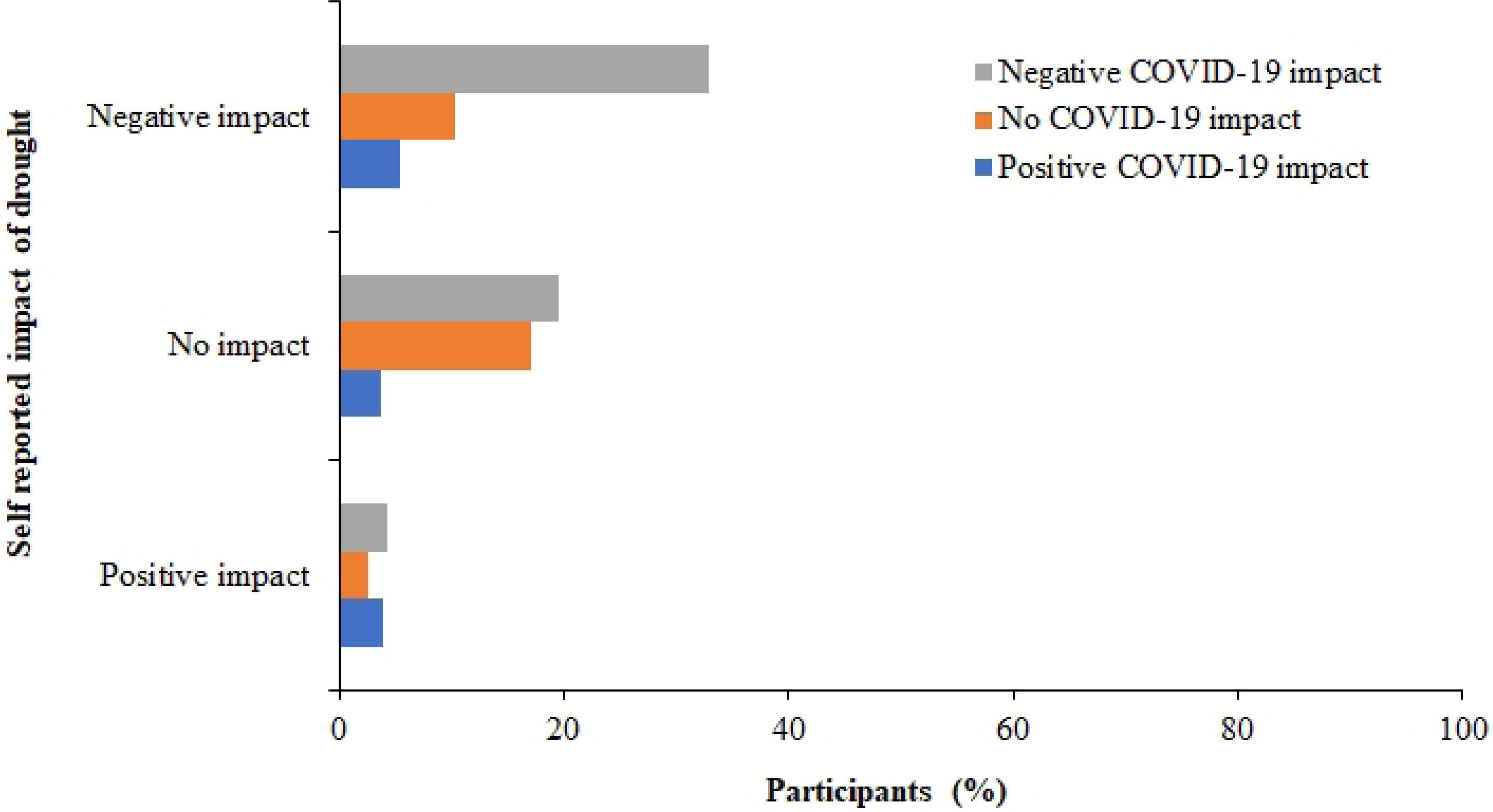
Self-reported impact of drought as a function of the perceived COVID-19 impact.

### Perceived Impact of COVID-19 and drought

Analysis of the respondents’ responses for the perceived impact of COVID-19 and drought showed that more than half of them (n=175, 56.8%) reported that COVID-19 had a negative impact on their lives (14.6% lost their income due to COVID-19), 13.6% (n=42) said it had a positive impact while the remaining 91 people (29.5%) were neutral. A slightly lower proportion of the respondents stated that they were negatively impacted by the drought (n=145, 48.7%), 11.1% (n=33) reported a positive impact while the rest were neutral (n=120, 40.2%).

### Lifestyle Behavioural Change due to COVID-19

All respondents (n=308) answered questions regarding the impact of COVID-19 on lifestyle factors (smoking, nutrition, alcohol, physical activity and sleep) and their responses are presented in Figure 1. The figure shows that, more than half of the respondents (56.4%) were active drinkers and over one quarter (26.3%) were active smokers. Across all lifestyle factors, the majority of the respondents reported no change in any of the measured lifestyle factors due to COVID-19. However, a few people (17.8% and 12.2%, respectively) reported increase in alcohol intake and smoking due to the pandemic. The greatest impact of the COVID-19 was observed among those who became inactive due to the pandemic. Compared with those who reported that they had more exercise due to the pandemic, those that reported less exercise were higher by 21.1%. The COVID-19 pandemic had more a positive than a negative impact on the respondents’ diet intake, but sleep patterns were adversely affected in a quarter of the respondents in this study.

### Relationship between psychological distress, lifestyle and disposition of the respondents

Current lifestyle behaviour at the time of this survey alongside distress levels is summarised in Table 2, Figure 6. The mean GLS score was 11.3 ± 1.63 (range, 7 – 15) and was significantly negatively correlated with psychological distress (r=-0.27, *P*<0.001), such that those with a higher positive lifestyle score reported lower scores for psychological distress. As shown in figure 2, the K-5 scores for psychological distress were significantly higher in respondents who slept for 7 hours or less a night, compared to those who slept more than 6 hrs a night (*P*<0.001). Similarly, a statistically significant correlation was observed between psychological distress scores and smoking (p=0.009) with *post-hoc* test revealing that heavy smokers had significantly higher psychological distress scores than non-smokers. No significant correlation was found between psychological distress scores and the other lifestyle variables of exercise, nutrition and alcohol intake.

### Association between Psychological Distress and Change in Lifestyle Behaviours, Income Loss and Greater Perceived COVID-19 Impact and Hopefulness

Table 3 presents the mean psychological distress scores in relation to the changes in lifestyle and income factors due to COVID-19. From the table, psychological distress scores varied significantly with changes in all lifestyle behavioural factors except for alcohol consumption. The scores were significantly higher in those who reported increase in smoking than those who reported no change (*P*<0.001). Respondents who reported either increased or decreased sleep, people who reported less exercise, and people who smoked more reported significantly higher scores for psychological distress than those who reported no change in these behaviours. Surprisingly, those who reported improvement in their nutrition had significantly higher scores for psychological distress than those with no change to their nutrition.

**Table 3:** Lifestyle Behaviours Correlated with Distress at Time of Survey. Significant *P*-values are bolded. Independent samples t-test was used for analysis of the sleep variable. One metabolic equivalent task (MET) is defined as energy expenditure at rest per minute, 500 MET is equivalent to 150 minutes of moderate physical activity (at approximately 3.33 MET) or 75 minutes vigorous activity (at approximately 6.66 <u>MET).</u>

| Lifestyle Factor | N (%) | K-5 mean (±SD) | F score | df | *P*-value |
| --- | --- | --- | --- | --- | --- |
| **Smoking** |  |  |  |  |  |
| Heavy (>5 cigarettes/day) | 36 (11.7) | 12.8 (5.2) | 4.800 | 2 | **0.009** |
| Moderate (<5 cigarettes/day) | 3 (1.0) | 10.7 (30.1) |  |  |  |
| Non-smoker | 269 (87.3) | 10.2 (4.4) |  |  |  |
| **Nutrition** |  |  |  |  |  |
| Does not meet 5 veg, 2 fruit | 42 (13.6) | 9.9 (4.6) | 0.830 | 2 | 0.437 |
| Meets either 5 veg or 2 fruit | 258 (83.8) | 10.6 (4.5) |  |  |  |
| Meets both 5 veg, 2 fruit | 8 (2.6) | 9.1 (4.5) |  |  |  |
| **Alcohol Intake** |  |  |  |  |  |
| High AUDIT-C | 10 (3.2) | 12.8 (4.5) | 1.505 | 2 | 0.224 |
| Moderate AUDIT-C | 52 (16.9) | 10.7 (4.2) |  |  |  |
| Low AUDIT-C | 246 (79.9) | 10.3 (4.6) |  |  |  |
| **Exercise** |  |  |  |  |  |
| MET score <500 | 175 (56.8) | 10.7 (4.5) | 2.326 | 2 | 0.1 |
| MET score 500-1000 | 66 (21.4) | 11.0 (4.6) |  |  |  |
| MET score >1000 | 67 (21.8) | 9.4 (4.4) |  |  |  |
| **Sleep!** |  |  |  |  |  |
| <7 hours | 118 (38.3) | 11.8 (5.1) | 15.957 | 1 | <0.001 |
| >7 hours | 190 (61.7) | 9.7 (4.0) |  |  |  |

Respondents whose incomes were reduced had significantly higher psychological distress scores than those whose incomes were increased or unaffected, during the COVID-19 pandemic. Similarly, those who reported a negative COVID-19 impact were also significantly more psychologically distressed than the other groups (see Table 4).

**Table 4:** Differences in mean psychological distress scores (K5) with the changes in lifestyle behavioural and income due to COVID-19.

| Variables | N (%) | Mean(±SD) | F value | P-value |
| --- | --- | --- | --- | --- |
| SNAPS Factor |  |  |  |  |
| Smoking (n=73) |  |  |  |  |
| Less smoking | 8 (11.0) | 10.3 (4.0) | 10.79 | <0.001 |
| No change | 47 (64.4) | 9.8 (3.8) |  |  |
| More smoking | 18 (24.7) | 14.9 (4.5) |  |  |
| Nutrition (n=286) |  |  |  |  |
| Worse nutrition | 41 (14.3) | 11.6 (4.1) | 5.22 | 0.006 |
| No change | 224 (78.3) | 10.0 (4.5) |  |  |
| Better nutrition | 21 (7.3) | 12.8 (4.9) |  |  |
| Alcohol (n=165) |  |  |  |  |
| Less alcohol | 22 (13.3) | 11.1 (4.8) | 1.94 | 0.148 |
| No change | 116 (70.3) | 10.4 (4.6) |  |  |
| More alcohol | 27 (16.4) | 12.2 (4.0) |  |  |
| **Exercise (n=283)** |  |  |  |  |
| Less exercise | 97 (34.3) | 11.4 (4.4) | 3.83 | 0.023 |
| No change | 148 (52.3) | 9.8 (4.4) |  |  |
| More exercise | 38 (13.4) | 10.7 (5.0) |  |  |
| **Sleep (n=286)** |  |  |  |  |
| Less sleep | 66 (23.1) | 13.2 (4.4) | 26.31 | <0.001 |
| No change | 202 (70.6) | 9.3 (3.9) |  |  |
| More sleep | 18 (6.3) | 13.3 (5.9) |  |  |
| **Income changes due to COVID-19** |  |  |  |  |
| Income reduced | 43 (15.0) | 12.8 (5.1) | 7.55 | <0.001 |
| No change in income | 210 (73.4) | 10.1 (4.4) |  |  |
| Income increased | 33 (11.5) | 9.5 (4.0) |  |  |
| **Perceived COVID-19 impact** |  |  |  |  |
| Negative COVID impact | 161 (56.3) | 11.7 (4.7) | 15.23 | <0.001 |
| Neutral COVID impact | 86 (30.1) | 8.9 (3.7) |  |  |
| Positive COVID impact | 39 (13.6) | 8.7 (4.0) |  |  |

There was a significant correlation between having a negative disposition (low THS) and higher psychological distress scores among the respondents (r = -0.365, *P*<0.001). One way ANOVA revealed that those who perceived a negative or no impact of COVID-19 had significantly lower mean THS than those who perceived a positive impact of COVID-19 (43.4 ± 10.4 and 43.0 ± 11.6 vs 48.9 ± 8.9, *P*=0.008) in this study.

### Association Between Subjective COVID-19 Impact and Lifestyle Change, Disposition, Income Loss and Drought Impact

As shown in Supplementary file (S1 Table 1), there was no significant correlation between self-reported COVID-19 impact and gender (F=2.059, p=0.357), age (F=6.966, p=0.138) or occupation (F=3.586, p=0.733). Current lifestyle was not significantly correlated with perceived COVID-19 impact, as measured in both individual lifestyle factors (S1 Table 2) and the composite GLS (F=1.817, *P*=0.164).

There was a significant association between subjective COVID-19 impact and some changes in lifestyle, specifically with increased smoking and decrease in hours of sleep (Table 5). Those who reported increased smoking and either more or less sleep were significantly more negatively impacted by COVID-19 than those who reported no changes. Notably, the impact of COVID-19 varied with the respondents’ change in income (χ*2* = 21.80, p=0.005), such that a greater degree of income loss was associated with a higher self-reported negative impact of COVID-19.

**Table 5:** Perceived COVID-19 Impact correlated with Changes in Lifestyle Behaviour.

| Variables | Negative COVID-19 impact | No impact | Positive COVID-19 impact | *P*-value |
| --- | --- | --- | --- | --- |
| Lifestyle Factors |  |  |  |  |
| Smoking |  |  |  |  |
| More smoking | 14 (66.7) | 4 (19.0) | 3 (14.3) | 0.027 |
| About the same | 27 (52.9) | 13 (25.5) | 11 (21.6) |  |
| Less smoking | 6 (60.0) | 0 (0.0) | 4 (40.0%) |  |
| Nutrition |  |  |  |  |
| Worse nutrition | 28 (63.6) | 8 (18.2) | 8 (18.2) | 0.395 |
| No change | 134 (55.1) | 78 (32.1) | 31 (12.8) |  |
| Better nutrition | 13 (61.9) | 5 (23.8) | 3 (14.3) |  |
| **Alcohol** |  |  |  |  |
| More alcohol | 21 (67.7) | 9 (29.0) | 1 (3.2) | 0.103 |
| No change | 63 (52.5) | 42 (35.0) | 15 (12.5) |  |
| Less alcohol | 12 (52.2) | 5 (21.7) | 6 (26.1) |  |
| **Physical Exercise** |  |  |  |  |
| Less exercise | 70 (67.3) | 25 (24.0) | 9 (8.7) | 0.072 |
| No change | 83 (51.2) | 54 (33.3) | 25 (15.4) |  |
| More exercise | 20 (51.3) | 11 (28.2) | 8 (20.5) |  |
| **Sleep** |  |  |  |  |
| Less sleep | 51 (68.0) | 18 (24.0) | 6 (8.0) | 0.005 |
| No change | 110 (51.2) | 73 (34.0) | 32 (14.9) |  |
| More sleep | 14 (77.8) | 0 (0.0) | 4 (22.2) |  |
| **Income change** |  |  |  |  |
| Significantly lower | 20 (83.3) | 3 (12.5) | 1 (4.2) | 0.005 |
| Slightly lower | 17 (77.3) | 4 (18.2) | 1 (4.5) |  |
| About the same | 124 (55.6) | 74 (32.5) | 29 (12.8) |  |
| Slightly greater | 11 (39.3) | 8 (28.6) | 9 (32.1) |  |
| Significantly greater | 3 (42.9) | 2 (28.6) | 2 (28.6) |  |

Figure 3 presents the perceived impact of the drought as a function of the COVID-19 impact, among the respondents. People’s perception of the drought impact was found to be significantly associated with how they were impacted by the COVID-19 pandemic (χ*2* = 31.93, *P*=0.005). Those who reported a negative impact of COVID-19 were significantly more likely to perceive that the drought had affected them adversely, and vice versa.

### Qualitative Data

Some common themes relating to the impact of COVID-19 on lifestyle factors (Table 6) were identified, with general impacts salient throughout including a predominantly negative impact on relationships, socialisation, and the ability to freely participate in in-person events. Respondents identified a reduced sense of connection and support, which was occasionally related to increased feelings of depression. Others also identified an increased level of stress and anxiety due to work or COVID-19 requirements. Most respondents did not smoke; however, COVID-19 did provide one individual with the impetus to quit; one participant reported relapsing during COVID-19, but this may or not have been a causative relationship, and no respondents described taking up smoking. Most respondents felt that COVID-19 had a negative impact on their ability to exercise, with the most significant limiting factors including fear of going out and closures/cancellations of facilities such as the gyms. Conversely, some respondents attributed increased levels of exercise to increased time due to less work and fewer social commitments.

**Table 6:** Qualitative Survey Responses for Lifestyle Factors.

| Topic | Quotes |
| --- | --- |
| **General** | • *“Being a single parent, I found not being able to visit friends and family or receive visitors in my home to be extremely isolating. Having no adult face to face contact messed with my mental health.”*<br>• *“Stress from COVID, winter, family...has made me depressed and has led to lethargy.”*<br>• *“Becoming more isolated”*<br>• *“Visitors from coast don’t visit as often”*<br>• *“Mental health... anxiety, lots of rules etc. Stress as a business owner.”* |
| **Smoking** | • *“2 weeks into quitting and have reduced by half. COVID has had a part in stronger motivation to quit.”*<br>• *“I was trying to quit but gave up trying when the panic was declared.”* |
| **Alcohol** | • *“[I drink] more due to work stress than COVID but COVID created the extra work stress”*<br>• *“Since the pubs closed I feel like I have not been able to get my drinking under control”*<br>• *“I am a social drinker so COVID has made me drink substantially less”*<br>• *“Drinking alcohol made [stress] worse, so I stopped [drinking] in June* |
| **Exercise** | • *“I am avoiding the gym because I can’t risk catching COVID and passing it on to elderly people”*<br>• *“Gyms either closing or having reduced operating hours impacts my exercise regime”*<br>• *“My year of planned running events was cancelled.”*<br>• *“With working from home during the lockdown, I was able to fit more exercise into my day.”* |
| **Sleep** | • *“I can't switch off”*<br>• *“I wake up through the night worrying about the future”*<br>• *“If everyone complied... life will be able to move forward”*<br>• *“During the first lockdown I had trouble sleeping ... but now I’m ok”*<br>• *“Other issues are causing sleep deprivation”*<br>• *“I have young children”* |

The impact of COVID-19 on alcohol consumption tended to follow two separate trends. Some regular alcohol consumers reported an increase in their alcohol intake due to stress, whereas social drinkers tended to consume less alcohol due to fewer social events. The impact of COVID-19 on sleep habits also showed effects in both directions, with some reporting less sleep due to anxiety regarding health, work, global events and young children, while others reported longer, better sleep and more naps, whilst working from home.

The effects of COVID-19 on general mental health of the respondents, appeared to be related to the increase in isolation and stress resulting from increased work demands. The effects of COVID-19 on lifestyle behaviours were variable and dependent on the individual’s circumstances.

## Discussion

This study examined the association between lifestyle behaviours (smoking, nutrition, alcohol, physical activity and sleep), lifestyle behavioural change during COVID-19, psychological distress and the reported perceived impact of COVID-19 in a rural Australian population, at a time of significant drought. Key findings were that COVID-19 has, on average, had a negative effect on all domains of lifestyle in this rural community. Those with the poorest lifestyles reported the highest levels of psychological distress during the pandemic, particularly less sleep and increased smoking. In terms of lifestyle change, a negative change in lifestyle (less sleep, poorer nutrition, increased smoking, and less exercise) was associated with an increase in psychological distress, which was consistent with the study’s hypothesis. Interestingly, the respondents who reported an improvement in their lifestyle during the lockdown, had similarly elevated psychological distress scores compared to those whose lifestyle remained unchanged. Those who felt the most affected by the COVID-19 pandemic, were also more likely to be distressed and this appears to overlap with those who felt most impacted by the drought before the pandemic. Notably, loss in income was significantly correlated with both higher psychological distress and greater self-reported COVID-19 impact in this study.

Several reports outlined earlier have found that, COVID-19 has had a significant impact on lifestyle behaviours and our study confirms this in this rural population. We found that a significant proportion of the respondents reported changes in each lifestyle domain, ranging from 21% with nutrition to 46% with exercise, with more people reporting negative than positive change across all domains. Exercise was also the most impacted domain in Stanton et al.’s cohort in April 2020, during the COVID-19 lockdown, with 69% of their respondents reporting an impact(6).

Given the well-established relationships between all lifestyle domains that we assessed and mental health, it was expected that both poor current lifestyle and the adoption of poor lifestyle habits during COVID-19 to correlate with higher psychological distress, and vice versa. However, inadequate sleep and active smoking were the only lifestyle behaviours that significantly correlated with psychological distress. Lack of correlation with other well-known factors like exercise may reflect unique characteristics of the rural sample or underpowering of our study resulting in weaker associations. Similar to our findings, Stanton et al (7). found that sleep had the strongest correlation with depression out of all lifestyle factors. Sleep is strongly associated bi-directionally with depression, with sleep duration and architecture being disrupted by depression and, concomitantly, sleep deprivation being a major risk factor for developing depression(7). The COVID pandemic has been associated with poor sleep widespread among the general population, with healthcare workers strongly affected(25). Regarding smoking, depression and psychological distress is known to predict smoking, likely representing ‘self-medication’ and shared underlying environmental causes, with chronic smoking causing neurophysiological changes that promote depression(26). Subsequently, when pooled into our composite GLS, we found that poorer lifestyles were associated with higher levels of distress during the COVID-19 pandemic, confirming our hypothesis and suggesting the potential utility of this global measure in providing an overall lifestyle assessment.

Regarding the change in lifestyle behaviours adopted due to COVID-19, we found a negative change in all the domains of the respondents’ lifestyle (less sleep, poorer nutrition, increased smoking, and less exercise), with the notable exception of impact on alcohol consumption. These were associated with higher psychological distress. Ingram et al.’s UK cohort also had no correlation between change in alcohol consumption and negative mood status unlike other lifestyle risk factors; they hypothesised that this may reflect the positive effect of alcohol on peoples mood, under certain social circumstances(8). Furthermore, Hu et al. found that, during the COVID pandemic, a decreased vegetable, fruit, and breakfast intake was associated with lower subjective wellbeing(27). Concerning the unexpectedly increase in psychological distress found among those who adopted healthier lifestyle behaviours, one hypothesis was a “self-medication” theory where those with high distress perhaps attempted to improve their anxiety through healthy outlets. Another theory borne out of the literature suggests that the process of adopting and maintaining a healthier lifestyle may in itself cause distress(28).

It was also found that a decrease in income was associated with an increase in psychological distress at the time and a greater negative self-reported COVID impact. Economic recessions have been shown to have a devastating effect on mental health. At the time of the current survey, over 206,000 people were unemployed due to COVID-19(29). A recent large prospective, longitudinal study of over 34,000 respondents found that participants who reported a decrease in income over a three year period, were 30% more likely to report a mental health or substance disorder(10). Furthermore, significant financial stress has been associated with increased interpersonal stressors, greater psychological distress and lower levels of psychological well-being(30). It is suggested that economic measures such as JobKeeper and JobSeeker(31) may have prevented the worst of the impact as government policy directed towards financial protection of Australians during COVID has been shown to improve mental health outcomes(32).

Regarding the perceived COVID-19 impact, respondents were asked to rate the severity of COVID-19’s impact on their life in general. This question was intended to capture the perceived overall psychological burden of the pandemic, including the effects of mandated restrictions on lifestyle, work, and education, broader social changes and fear of contracting the disease itself.

Change in sleep and smoking habits were significantly correlated with the respondents’ perceived COVID-19 impact, which was expected, though it is notable that the change in other lifestyle behaviours during the pandemic were not significantly correlated with the perceived impact. This study demonstrated that current lifestyle did not correlate with perceived COVID impact, either as individual behaviours or as a composite GLS. As such, respondents with poorer and healthier lifestyles reported similar impact of COVID-19; however, those who reported a negative change in their sleep and smoking behaviours during COVID-19 pandemic, reported an increase in the impact of COVID-19. This lack of relationship between individuals’ current lifestyle and their perception of COVID-19 impact suggests that other factors such as income decline (discussed above) appear to be more dominant; nonetheless, as poor lifestyle continues to correlate with overall psychological distress, it remains important for mental health even in the context of additional stressors.

Respondents with a high level of self-reported COVID-19 impact also reported higher psychological distress. Considering that our result did not find any correlation between distress and a negative disposition, their report of higher distress can be attributed to the impact of COVID on their lives, rather than their disposition. Consistent with the hypothesis that COVID-19 may have directly caused poorer mental health outcomes, one large longitudinal study has found that people without previous mental health disorders reported an increase in symptom severity on scales used to measure mental health when compared to pre-pandemic levels(19).

The current study found no significant association between demographic factors and COVID-19 impact with similar self-reported COVID-19 impacts across age, sex and occupation. A large systematic review on the impact of COVID-19 on mental health identified women, students, age <40, pre-existing psychiatric conditions and increased exposure to social media as risk factors for increased distress, during the pandemic(20). Similarly, the most vulnerable people who had lost their jobs, lived alone or were living in poorly-resourced areas, were providing care to dependent family members of marginalised minorities, women or young people, had the most severe impact(21).

The association between perceived COVID-19 impact and drought impact within the same population suggests that there are common mechanisms or vulnerabilities that may impact rural populations, such as the impact of external stressors on income. Throughout the drought, farmers in NSW experienced significant distress due to the effects of the drought on themselves, their families and their communities(17). Specifically, farmers who experienced financial hardship or were isolated from their communities by virtue of working in remote areas, were prone to drought related stress(17). The results of this study suggest that these other risk factors may have been compounded by the COVID-19 pandemic in 2020 resulting in the association between drought-related stress and self-reported COVID-19 impact.

### Limitations and strengths

The limitations of the current study include the relatively small sample size, predominantly composed of respondents from one rural town in Australia with access to the Internet on a smartphone or computer. The data was largely skewed towards older female respondents, who are not representative of the entire rural Australian population. This limitation existed due to the inability to distribute the survey in person due to the pandemic and because the online groups where the surveys were distributed, had a relatively inactive younger population and included only individuals who had access to the Internet on a smartphone or computer. Furthermore, the study design did not differentiate between potentially important underlying social and demographic subgroups such as parental status. Glenister et al. reported an increase in alcohol only among women who were living with children at home as opposed to those without(9), and the effect of alcohol on mood is known to vary between social situations. Not taking these subgroups into account may have confounded the correlations found between lifestyle behaviours and COVID-19 impact, by obscuring the underlying correlations in opposite directions. A further limitation of the study was the reliance on self-reported data that cannot be independently verified. The data may therefore have been incorrectly recalled or exaggerated. Cross-sectional studies such as the current one make it difficult to delineate cause and effect. For example, lifestyle change, and COVID-19 impact may work bidirectionally to influence each other. Further research involving alternative data gathering methods and perhaps using a longitudinal study are needed to address these limitations. The current study developed a GLS based on evidence-based recommendations in each of the 5 key lifestyle domains recommended by the RACGPs to be included in standardised lifestyle screening. Although, this scoring system has not, as yet, been validated beyond the current study for use in measuring overall lifestyle, it could potentially be refined into a clinically applicable scoring tool to be used in practice. Further validation is required to enhance its utility inpredicting other established lifestyle-dependent conditions, such as cardiovascular disease and diabetes – i.e. before it can be formally used as a GLS. Future research into the impact of COVID-19 on young Australians in a rural setting should consider this.

### Future directions for research

As those with perceived increased drought impact were more likely to report increased COVID-19 impact, a direction for future research is the extent to which drought relief initiatives may build resilience within communities to prepare them for future crises. The relationship between previous crises such as drought and the COVID-19 pandemic may also be informative. To the best of the authors’ knowledge, a GLS applicable in the clinical setting has never been developed or widely accepted. The proposed GLS is based on lifestyle recommendations from relevant leading health organisations and has been shown to correlate well with high levels of distress in this study. Further research into whether this clinical tool can be validated for further mental and physiological health conditions is required. Several measured lifestyle factors demonstrated correlational trends in opposite directions – i.e. in both positive and negative directions. More in-depth demographic studies are needed to elucidate factors that may contribute to one or other trend, which in turn may provide further information in relation to those who are most vulnerable to distress.

### Community Engagement

Due to COVID-19 and the difficulty in distributing surveys in-person, community engagement was primarily achieved online by contacting local councils, who distributed a link to the survey through email, and additionally by posting on Facebook groups containing members living in the Central West of NSW. Reminders to complete the survey were posted on these groups to increase response numbers. The survey medium and mode of recruitment highlight a likely shift in the context of COVID-19 and, in future, towards use of IT and community engagement online. These surveys can be completed on mobile devices or on a computer, highlighting the multi-modal and ease of access of these on-line surveys, irrespective of the physical location of the participant.

## Conclusion

In conclusion, the current study suggests that rural Australians’ lifestyle parameters such as smoking, sleeping, nutrition, exercise but not alcohol, were worse, during the COVID-19 pandemic. Greater COVID-19 impact was associated with higher distress and a greater change in overall lifestyle. These findings present important implications for health professionals towards a greater understanding of COVID-19 and its effect on lifestyle and mental health amongst patients and directing their treatment strategies accordingly. As a contextual factor that continues to evolve at the time of writing, the perceived impact of COVID-19 on the lifestyle factors under consideration is evolving and further research is needed to investigate the clinical utility of these lifestyle behaviours.

## Data Availability

All relevant data are within the manuscript and its Supporting Information files.

## Conflict of interest

There are no conflicts of interest to declare. The authors received no external funding.

## Authors’ Contributions

JC, KC, JG, HX, RV, JB designed the study and developed study materials. JC, KC, JG, HX recruited study respondents. JC, KC, JG, HX, RV, JB drafted the manuscript. Data analysis was conducted by JC, KC, JG, HX, with input from JB. All authors contributed to writing, editing and approval of the final manuscript.

## Acknowledgements

We have no acknowledgements to make.

## Ethics Approval and Consent to Participate

Ethics approval was obtained from Western Sydney University Human Research Ethics Committee (H13796). All respondents were provided with written informed consent prior to participation. The paper contains no identifying information. The study protocol conformed with the tenets of the declaration of Helsinki for studies involving human respondents.

## Notes

### Competing Interest Statement

The authors have declared no competing interest.

### Funding Statement

The author(s) received no specific funding for this work.

### Author Declarations

Ethics approval was obtained from Western Sydney University Human Research Ethics Committee (H13796).

